# Life without sex: Large-scale study links sexlessness to physical, cognitive, and personality traits, socioecological factors, and DNA

**DOI:** 10.1101/2024.07.24.24310943

**Authors:** Abdel Abdellaoui, Laura W. Wesseldijk, Scott D. Gordon, Joëlle A. Pasman, Dirk J.A. Smit, Renáta Androvičová, Nicholas G. Martin, Fredrik Ullén, Miriam A. Mosing, Brendan P. Zietsch, Karin J.H. Verweij

**Author notes:** Corresponding author: Karin Verweij. Shared first authors (Abdel Abdellaoui, Laura W. Wesseldijk). Shared last authors (Brendan P. Zietsch, Karin J.H. Verweij).

## Abstract

Romantic (typically sexual) relationships are important to personal, physical, mental, social, and economic wellbeing, and to human evolution. Yet little is known about factors contributing to long-term lack of intimate relationships. We investigated phenotypic and genetic correlates of never having had sex in ∼400,000 UK residents aged 39 to 73 and ∼13,500 Australian residents aged 18 to 89. The strongest associations revealed that sexless individuals were more educated, less likely to use alcohol and smoke, more nervous, lonelier, and unhappier. Sexlessness was more strongly associated with physical characteristics (e.g. upper body strength) in men than in women. Sexless men tended to live in regions with fewer women, and sexlessness was more prevalent in regions with more income inequality. Common genetic variants explained 17% and 14% of variation in sexlessness in men and women, with a genetic correlation between sexes of 0.56. Polygenic scores predicted a range of related outcomes in the Australian dataset. Our findings uncover multifaceted correlates of human intimacy of evolutionary significance.

## Introduction

Understanding correlates of long-term sexlessness is of interest for at least two broad reasons. First, romantic partnerships, which are typically sexual, are often among the most important relationships in individuals’ lives, providing a host of personal, health, social, and economic benefits^1^. Those who go without may be vulnerable to, for example, poor mental health and loneliness, social embarrassment, and economic disadvantages (e.g. from lack of co- habitation), and those involved in online ‘incel’ (involuntary celibate) cultures are at risk of radicalisation.^2^ Understanding correlates of sexlessness may therefore inform strategies at the individual or societal level for removing barriers to finding fulfilling partnerships. Of course, this is not to say that sexual relationships are important to everyone – some 1% of the population reports no desire for sexual relationships (i.e. asexuality)^3^.

Second, long-term sexlessness may be a valuable trait with which to examine evolutionary hypotheses. The effort to understand the evolution of human nature generally involves understanding how selection pressures may have operated in the evolutionary past – that is, before widespread birth control and family planning. Because sex and reproduction are now largely separated in modern societies, reproductive success today is not a useful indicator of fitness as it would have manifested in the evolutionary past^4,5^. But lifetime number of mates (opposite-sex sexual partners) as an indicator of fitness also has drawbacks: notably, women’s reproductive success has always been limited by biology rather than mate quantity, and for both sexes mate quantity does not capture mate quality, which can affect longer-range reproductive success (i.e. fitness). However, these issues become moot if we consider the distinction between individuals who have no lifetime mates versus those who have at least one. Having no mates during the reproductive lifespan would have been an evolutionary dead end in any ancestral context. Many individuals today choose not to have children for reasons concerning the time, energy, and financial demands of child-rearing and fitting these demands with the conflicting demands of modern careers; but these contemporary constraints would not induce complete avoidance of sex. Therefore, lifetime sexlessness may be a better indicator of ancestral fitness and could serve as an important variable with which to supplement evolutionary analysis of selection.

As of yet, few studies have examined the demographic, individual, or environmental factors that characterise late-life virgins^6^. Eisenberg et al.^7^ (N = 7,589; age 25-45) and Ghaznavi et al. ^8^ (N = 11,553–17,850; age 18-39) examined characteristics that distinguished those who had never engaged in heterosexual intercourse. Eisenberg et al. ^7^ reported associations with greater religiosity, having a college degree, and lower drug and alcohol use, and Ghaznavi et al. ^8^ reported association with un- or underemployment, but there are several issues that make these findings hard to interpret. First, the focus on heterosexual sex makes the analyses heavily confounded by participants’ sexual orientation. Second, both samples consisted largely of younger adults, so it is unclear to what extent the findings apply to lifetime sexlessness as opposed to merely delayed onset of sexual behaviour. Third, only a limited range of characteristics were examined: demographic information and, in the case of Eisenberg et al. ^7^, self-reported health, drug and alcohol use, and body mass index. Fourth, while these samples were substantial in size, the low rate of lifelong sexlessness limited the statistical power to detect effects in these studies. Another study^9^ took a different approach to studying correlates of sexlessness and found that “incel” (involuntary celibate) posts on social media (Twitter) were more likely to originate from regions of the USA with male-biased sex ratio and higher income inequality. These findings suggest a link between sexlessness and local mating ecology; but the conclusion is tentative given the uncertain connection between the miniscule proportion (<.0001%) of posts with incel language and the substantial proportion of sexless individuals in the regions’ populations.

In the present study, we explore the correlates of sexlessness in data from the UK Biobank which consists of ∼500,000 middle and old age UK residents who have been genotyped and assessed with extensive testing and surveys^10^. Participants have reported their lifetime number of both opposite and same sex partners, so the analysis is less confounded by sexual orientation than previous studies. Participants are also largely past typical reproductive age; sexlessness at this stage of life (compared to that of the younger participants in previous studies) is more reflective of lifetime sexlessness, especially as it pertains to evolutionary analyses. Further, the dense phenotyping allows for a fuller characterisation of cognitive, health and other correlates of sexlessness, and the availability of genotype information enables us to estimate genetic associations of sexlessness with any trait that has been subject to genome-wide association study (GWAS), even if that trait is not assessed in the UK Biobank participants. Genetic correlations are especially useful in the context of evolutionary analyses, because these are more relevant to evolutionary responses to selection than are phenotypic correlations. We compare phenotypic and genetic correlates of sexlessness with those of childlessness to assess the degree to which sexlessness represents a distinct trait. We also investigate whether polygenic scores for sexlessness predict related outcomes in an independent Australian dataset. Last, the UK Biobank participants’ regions of residence and birth are recorded, enabling us to directly examine the association of regional sex ratio and income inequality with sexlessness.

## Results

### Phenotypic correlates of sexlessness and childlessness

Data on sexlessness were available for 405,117 British individuals of European descent, 218,744 females and 186,373 males. Of those, 3,929 individuals (2,068 females and 1,861 males, both ∼1%) responded that they have never had vaginal, oral, or anal intercourse. To examine phenotypic correlates of sexlessness, we selected 253 phenotypes (Table S1) that had an effective sample size larger than 10,000 and were related to domains of mental health, sleep, exercise, substance use, risk taking behavior, cognition, health, and occupation. Sexlessness was significantly associated with 149 of these traits (Table S2), of which 35 explained 1% or more of the variance (presented in Figure 1). The strongest associations were observed for phenotypes related to social connection and substance use. Sexlessness was associated with less and shorter mobile phone use, a lower chance of being in a confiding relationship, fewer friend and family visits, less alcohol and nicotine use, wearing glasses at an earlier age, and lower grip strength. Sexlessness was also associated with feeling more nervous and lonely and less happy, confirming its strong relevance to wellbeing.

**Figure 1:**
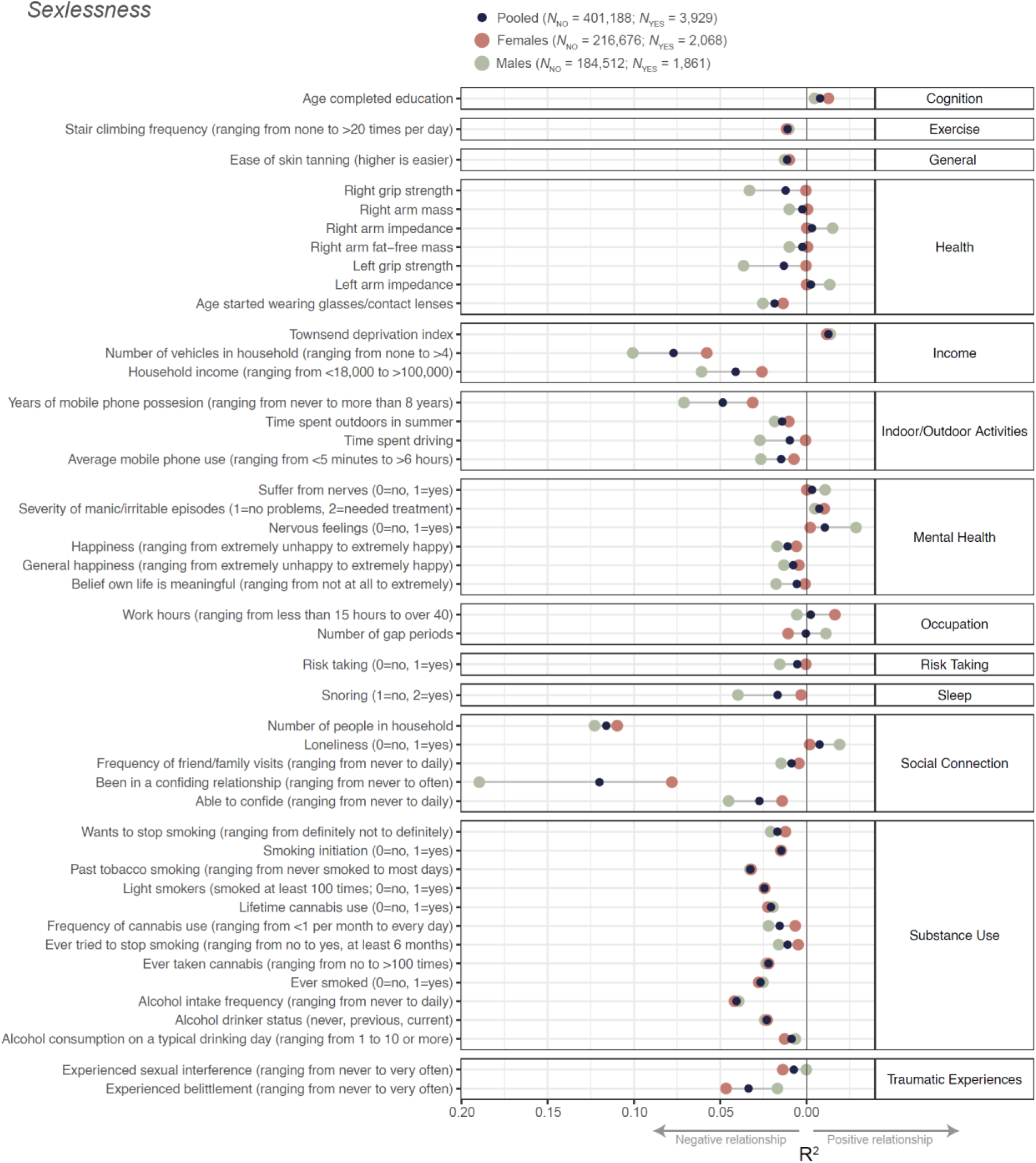
Phenotypic associations of sexlessness with health, psychological, and behavioural outcomes. Sexlessness was coded as 0=has had sex, 1=never had sex. Results are shown for the full (sexes pooled) sample, as well as for males and females separately. Only associations that were significant in at least one of the analyses (i.e. sexes pooled, males, or females) and explaining 1% or more variance are shown. Full results can be found in Table S2. Note that to improve clarity, in some instances variable names and coding have been changed from the original UK Biobank names/coding, see Table S1.

Data on childlessness were available for 455,979 individuals, of which 44,118 males reported to have not fathered any children and 46,251 females reported haven given birth to zero children (Ntotal = 86,464). A small subset of those (N = 1,847 males and 2,058 females) reported they had children, but did not have sexual intercourse in their life; we excluded these individuals from the analyses focused on childlessness, reducing the sample to 452,050 individuals. Childlessness was associated with 213 of the selected health, psychological, and behavioural outcomes, of which 10 traits explained 1% or more of the variance (Figure 2, Table S2). Childlessness was most strongly associated with phenotypes related to occupation, social connection, and sexuality. Furthermore, childlessness was less strongly associated with health, mental health, and substance use than was sexlessness. In line with the results for sexlessness, childlessness was also associated with fewer friend and family visits, a lower chance of being in a confiding relationship, believing that your life is less meaningful, feeling more nervous and lonely, and less happy, though to a lesser extent. In contrast, childlessness was also associated with working more hours, and it was not associated with any of the other health and mental health outcomes.

**Figure 2:**
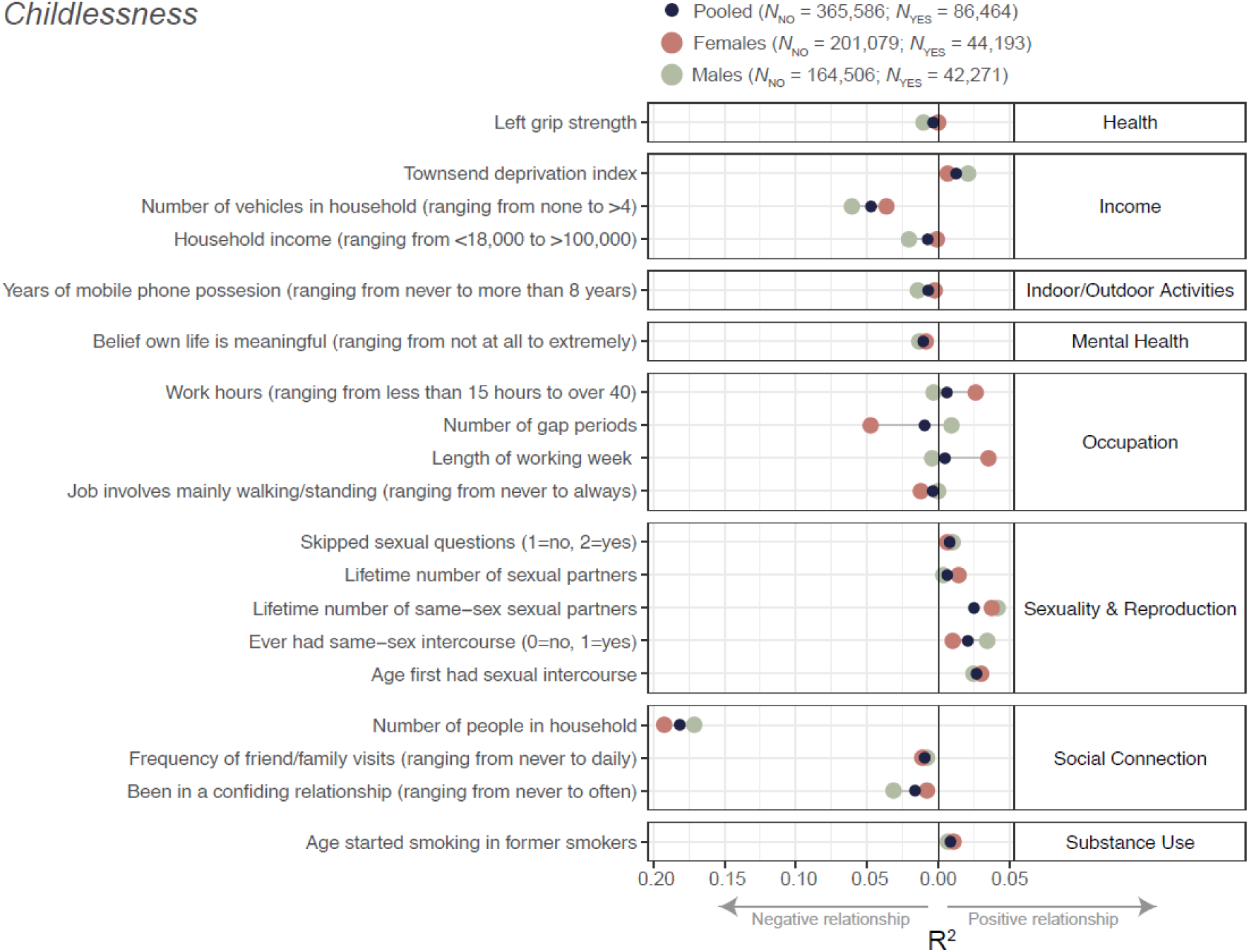
Phenotypic associations of childlessness with health, psychological, and behavioural outcomes. Childlessness was coded as 0=has children, 1=has no children. Results are shown for the full (sexes pooled) sample, as well as males and females separately. Only associations that were significant in at least one of the analyses (i.e. sexes pooled, males, or females) and explaining 1% or more variance are shown. Full results can be found in Table S2. Note that to improve clarity, in some instances variable names and coding have been changed from the original UK Biobank names/coding, see Table S1. Note that there were too few female participants who had nonzero numbers of ‘lifetime number of same-sex sexual partners’, so this estimate is missing.

Sex-specific analyses showed several differences between males and females in the correlates of sexlessness. Specifically, health factors such as grip strength and body measures, as well as income, snoring, mobile phone use, belief that one’s life is meaningful, being in a confiding relationship and being able to confide were more strongly associated with sexlessness in males than in females. Income and being in a confiding relationship were also more strongly associated with childlessness in males than in females, while occupational characteristics such as working hours were more strongly associated with childlessness in females than in males.

### Sexlessness in relation to regional sex ratio and income inequality

We investigated the associations between sexlessness and the sex-ratio of the region in which people were born or currently lived and between sexlessness and income inequality in these regions. The regional sex-ratios (% women) at the Middle Layer Super Output Area (MSOA) level were estimated from the census carried out by the Office of National Statistics in 2011. For regional income inequality we used the Gini coefficient, which was computed from the self-reported household income of the UK Biobank participants.

When considering birth place, there was no significant association between regional sex ratio and sexlessness (Figure 3). When considering current address, there was a small but significant negative association between regional sex ratio and sexlessness for males, whereby regions with relatively fewer females have higher rates of male sexlessness (*r* = -.07, *p* = .0004).

**Figure 3:**
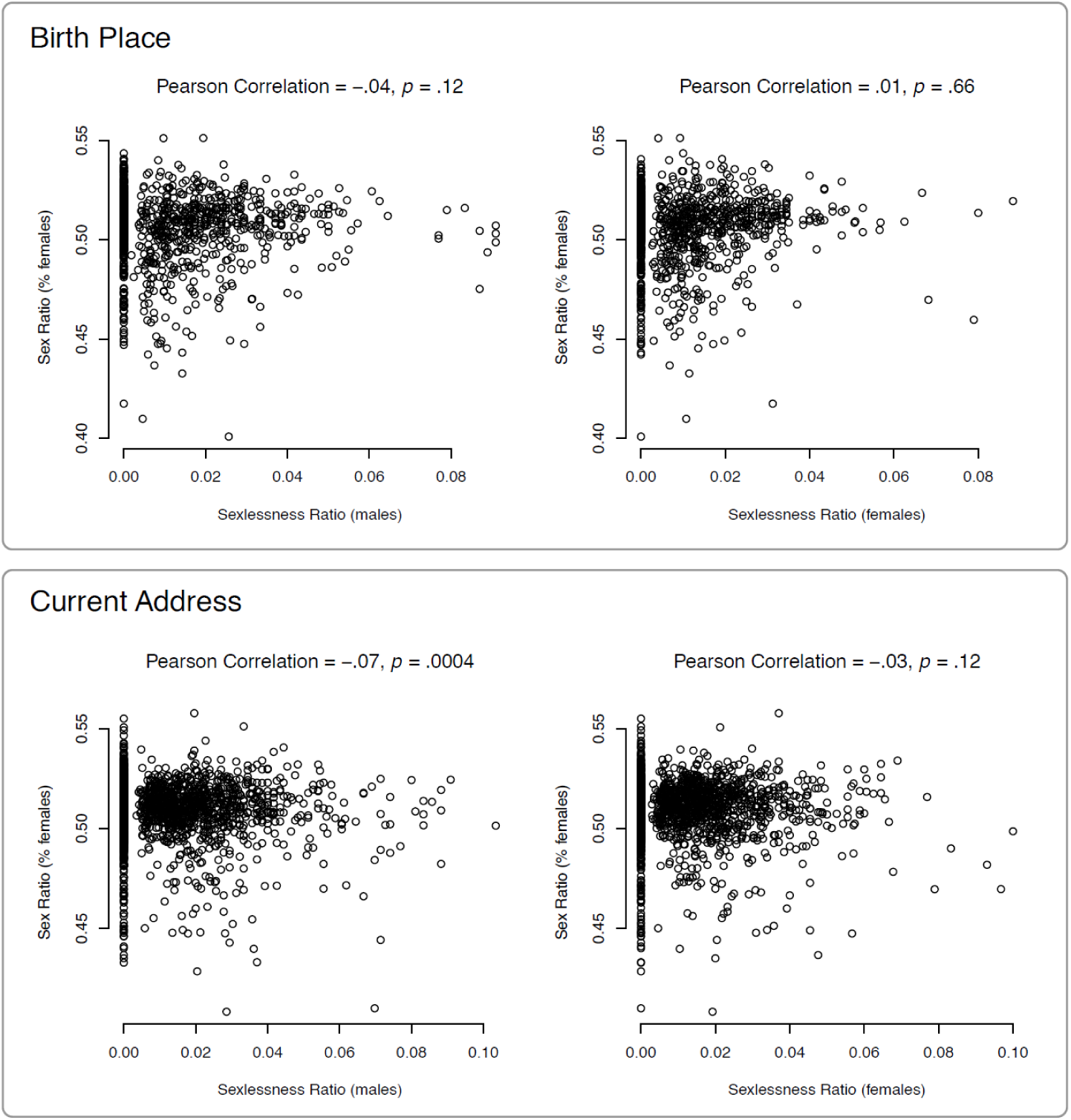
Sex ratio (proportion of women) versus sexlessness ratio, for birth place and current address. Sex ratio was based on census data and sexlessness ratio on UK Biobank data. Top panel: the Figures show results for 2,476 MSOA regions of the current address with at least 50 UK Biobank participants per region. The average sex ratio is .51. Bottom panel: The Figures show results for 1,334 MSOA regions of the birth place with at least 50 UK Biobank participants per region. The average sex ratio is .51. The average sexlessness ratio is .01 for males, and .01 for females.

The relationship between sexlessness and regional income inequality, as measured with the Gini coefficient, was significant and positive for current address (Figure 4) for males (*r* = 0.09, *p* = 2.8 × 10^-15^), females (*r* = 0.14, *p* = 3.6 × 10^-6^), and both sexes pooled (*r* = 0.15, *p* = 9.5 × 10^-13^), implying that sexlessness was associated with a higher regional income inequality. Again, there were no significant associations for birthplace.

**Figure 4:**
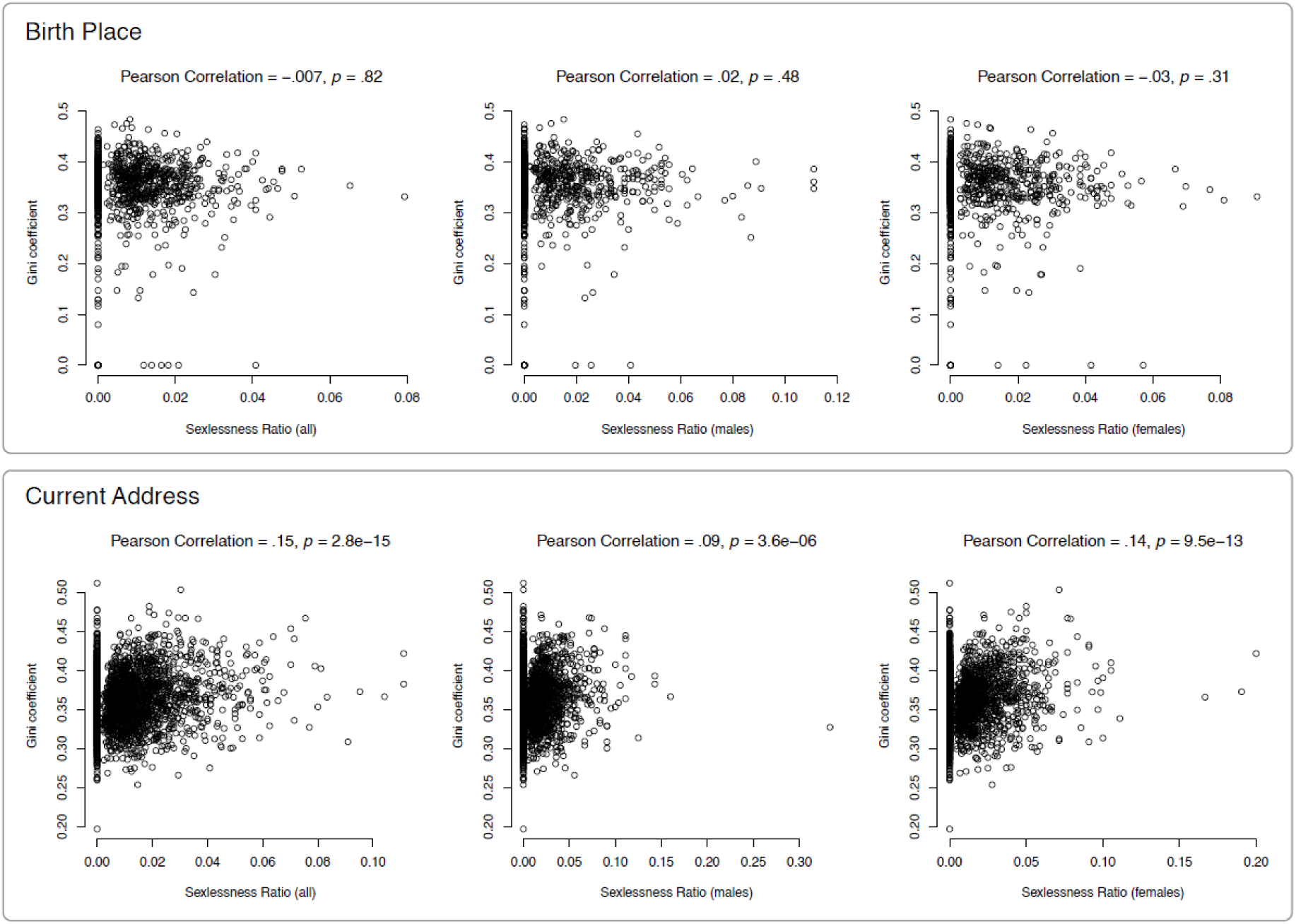
Income inequality versus sexlessness ratio for birth place and current address. Both income inequality and sexlessness ratio were derived from UK Biobank data. Top panel: the Figures show results for 1,499 MSOA regions of the birth place with at least 50 UK Biobank participants per region. The average GINI coefficient is .35. Bottom panel: The Figures show results for 2782 MSOA regions of the current address with at least 50 UK Biobank participants per region. The average GINI coefficient is .36. The average sexlessness ratio is .01 for the pooled sample, .01 for males, and .01 for females.

### Genome-Wide Association Study (GWAS)

GWAS analyses on sexlessness were conducted on 10.6 million single nucleotide polymorphisms (SNPs) in 404,470 UK Biobank participants of European descent, of which 3,897 reported to have been sexless (Table 1). There was one significant locus (top SNP rs912773) on chromosome 1 when pooling males and females (Figure 5). This locus is intergenic, with the closest gene LOC107984933 (see Figure S1 for a locus zoom plot). This SNP by itself had a miniscule effect size. In aggregate, though, SNPs across the genome were substantially associated with sexlessness: the SNP-based heritability of sexlessness as computed with LDSC regression^11^ accounted for 17% (SE = 4%, *p* = 2.3×10^-5^) and 14% (SE = 3%, *p* = 3.1×10^-6^) of the trait’s variance in males and females, respectively. Note that the combined SNP-based heritability (12%) is lower than the sex-specific heritability, likely because SNP-based heritabilities tend to decline as heterogenous groups are combined^12^. The genetic correlation between males and females is 0.56 (SE = 0.17, *p* = 0.0007), indicating that the SNP associations with sexlessness tend to be in the same direction in males and females, but only partially overlap. We used MAGMA to aggregate SNP effects at the gene level using positional annotations to compute gene-based *P*-values. None of the genes reached genome-wide significance at a Bonferroni corrected significance threshold of 2.67×10^-6^ (Figure S2), so we did not proceed with any further annotation of the association results. Please refer to Box 1 for some cautionary notes on the interpretation of genetic findings.

**Figure 5:**
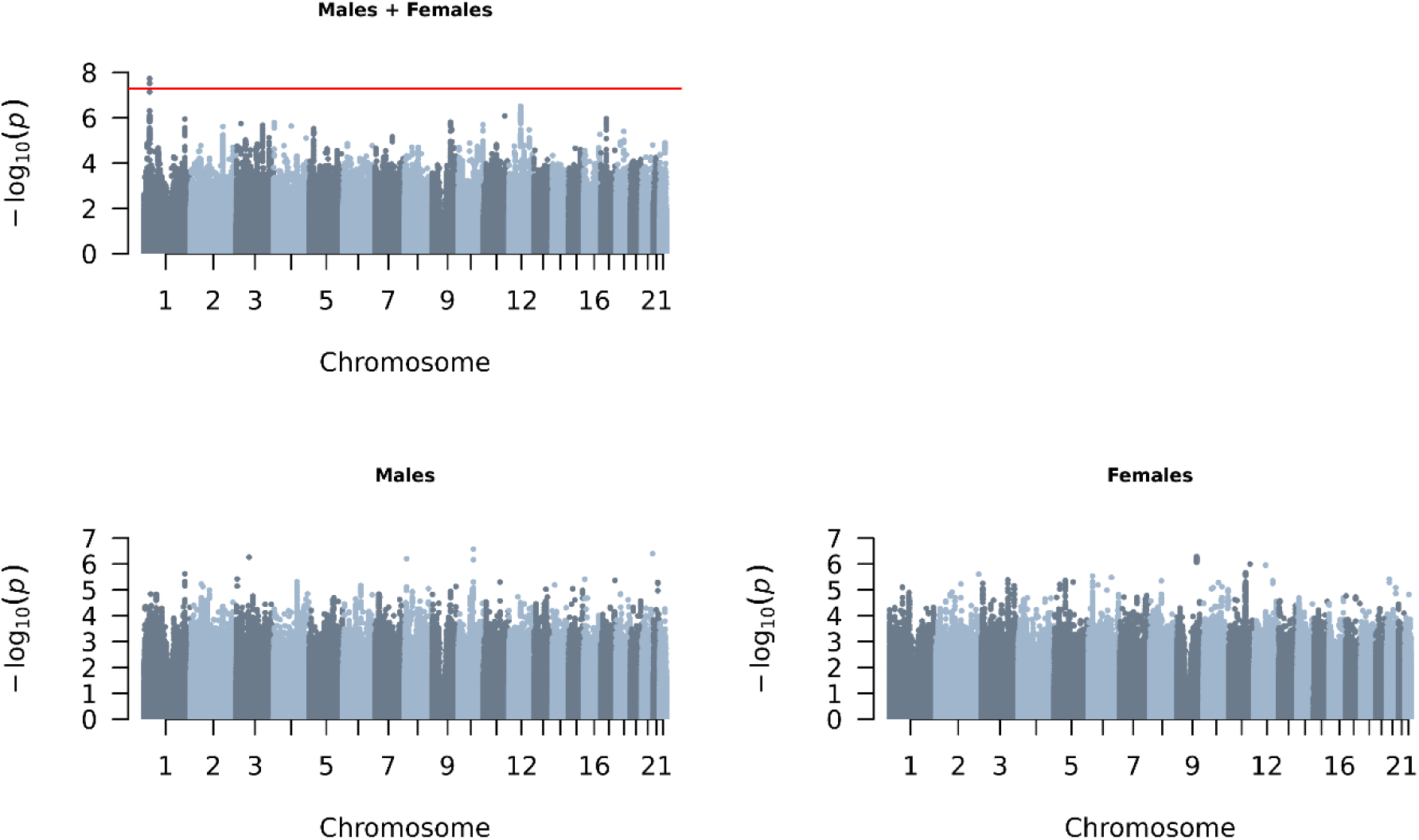
M**a**nhattan **plots of GWASs on sexlessness**. The x-axis shows 22 autosomes and the y-axis the –log10 P-values. The red line indicates the threshold of genome-wide significance (5E-08). The sample sizes and SNP-based heritabilities of these GWASs are displayed in Table 1.

**Table 1:**
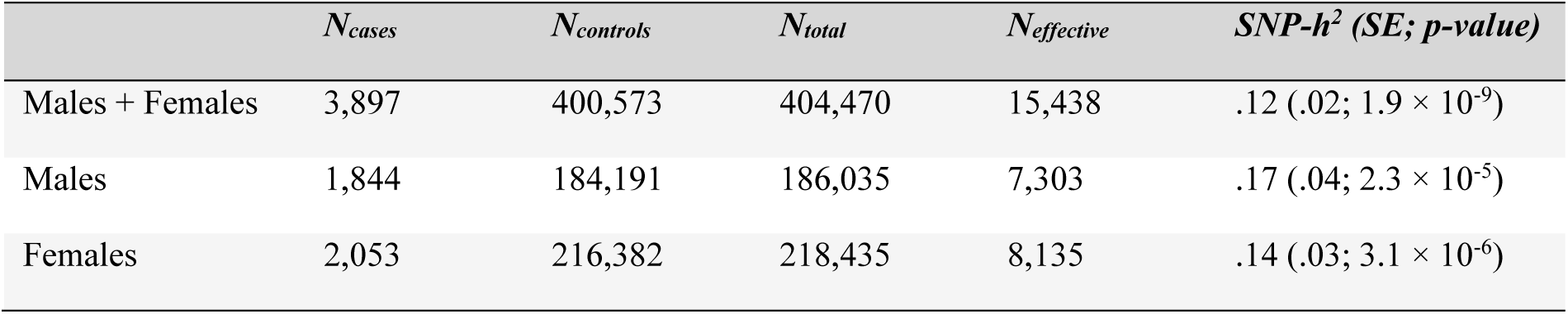
Sample sizes and SNP-based heritability (SNP-h^2^) of the sexlessness GWASs.

| | $N_{cases}$ | $N_{controls}$ | $N_{total}$ | $N_{effective}$ | $SNP-h^2$ (SE; $p$-value) |
| --- | --- | --- | --- | --- | --- |
| Males + Females | 3,897 | 400,573 | 404,470 | 15,438 | .12 (.02; $1.9 \times 10^{-9}$) |
| Males | 1,844 | 184,191 | 186,035 | 7,303 | .17 (.04; $2.3 \times 10^{-5}$) |
| Females | 2,053 | 216,382 | 218,435 | 8,135 | .14 (.03; $3.1 \times 10^{-6}$) |

**Box 1: Cautionary Notes**

Associations between genetics and behavior can be misinterpreted, sometimes with nefarious consequences. To minimize misinterpretations, we want to emphasize the complex nature of these associations. The genetic associations we discovered should not be presented as the discovery of “genes for late-life sexlessness” for several reasons outlined below.

Associations between genetic variants and behavioral traits likely reflect a culture-specific interplay between underlying heritable factors and environmental influences^13^. An association observed in a contemporary Western society should not be generalized as a universally applicable genetic effect on sexlessness. Therefore, our genetic and phenotypic associations may not apply to populations of different ancestries or cultural backgrounds.

Moreover, the polygenic scores derived from these genetic associations are not suited to predict individual-level outcomes regarding sexlessness. These polygenic scores are based on approximations of small effects within a large sample from a specific population. Therefore, it is inevitable that there will be many individuals who score high on such a polygenic score yet low on its target trait, and vice versa.

Overall, it is important to note that genetics is only one facet of the complex network of factors influencing behavior. Environmental, social, cultural, and individual factors all play significant roles, and their interactions with genetics further complicate the picture. This study points to possible avenues for investigation and should be seen as an exploration of correlations, not a claim of cause-and-effect relationships.

### Polygenic score analyses

We constructed polygenic scores from the GWAS results to validate their ability to predict sexlessness and related traits in independent samples, and to test for gene-environment correlations in the GWAS signal. Polygenic scores are (noisy) measures of an individual’s genetic predisposition for a trait, based on the sum of alleles weighted by their estimated effect on the trait of interest (here sexlessness).

In the UK-Biobank, we repeated the GWAS on sexlessness in a subset of the sample, excluding all siblings (and their relatives) and then created polygenic scores in the sibling sample. The polygenic scores significantly predicted sexlessness in this sample (Beta = 0.016 (SE = 0.005, *p* = 0.002)). We subsequently used these polygenic scores to test for gene-environment correlations at the family level for sexlessness. Gene-environment correlation occurs when individuals’ trait-relevant environment is associated with their genetic makeup. This could complicate our interpretation of the GWAS and polygenic score associations for sexlessness. To assess the extent to which our GWAS associations and polygenic scores for sexlessness could be explained by gene-environment correlation, we compared the prediction of sexlessness within and between families in the UK Biobank sample (see Methods for details). Adding the between-family effect to the model decreased the individual-level effect by only 7.4%, indicating the GWAS signal only captures gene-environment correlation effects to a very small degree. For comparison, in Selzam et al. (2019)^14^, the individual-level prediction for educational attainment decreased by 49%, IQ by 48%, BMI by 15%, and height by 12%. We therefore believe the GWAS associations and polygenic scores are minimally affected by gene-environment correlation, at least in terms of environments that differ between families.

To further validate the GWAS findings, we also examined the associations between polygenic scores for sexlessness and a number of relevant traits related to sexuality, mating, and attractiveness in an independent Australian sample. Polygenic scores for sexlessness were significantly associated with several phenotypes in the independent Australian target cohort (18-89 year olds, N ranged between 1,354 and 13,532) in the expected direction (Table 2, Table S3). The sexlessness polygenic score was significantly negatively associated with number of relationships of at least 3 months (for the sexes pooled), positively with never having had a sex partner (only for females), negatively with number of sex partners (sexes pooled), and positively with age at first intercourse (sexes pooled, males, and females). For some other variables (ever had a committed relationship, number of sex partners (males)), nominally significant associations (i.e. uncorrected for multiple testing) were found in the expected direction. That all of these associations are in the expected directions suggests the SNP associations discovered in the UK Biobank are capturing trait-relevant factors and not only artefacts of the UK population (e.g. ancestry/geography stratification). Little can be concluded from the non-significant associations, given the low power in this relatively small sample.

**Table 2.** Results of the polygenic score analyses for sexlessness in the independent Australian target sample.

| Phenotype | Sample | N | Slope | SE | z | p-value |
| --- | --- | --- | --- | --- | --- | --- |
| Current Partner (0=yes, 1=no) | Sexes pooled | 13532 | -0.372 | 3.802 | -0.098 | 0.922 |
|  | Males | 5396 | -2.008 | 5.626 | -0.357 | 0.721 |
|  | Females | 8136 | 4.044 | 5.905 | 0.685 | 0.494 |
| Ever had committed relationship (0=yes, 1=no) | Sexes pooled | 9501 | 7.956 | 2.988 | 2.663 | 0.008* |
|  | Males | 4005 | 5.662 | 4.334 | 1.307 | 0.191 |
|  | Females | 5496 | 8.271 | 4.414 | 1.874 | 0.061 |
| Number of relationships that lasted 3 months | Sexes pooled | 2466 | -63.319 | 20.270 | -3.124 | 0.002** |
|  | Males | 1015 | -18.167 | 27.822 | -0.653 | 0.514 |
|  | Females | 1451 | -81.030 | 31.429 | -2.578 | 0.010* |
| Having had sex partner (0=yes, 1=no) | Sexes pooled | 8516 | 3.493 | 1.794 | 1.947 | 0.052 |
|  | Males | 3602 | 4.354 | 2.657 | 1.639 | 0.101 |
|  | Females | 4914 | 9.874 | 3.064 | 3.223 | 0.001** |
| Number of sex partners | Sexes pooled | 2309 | -77.102 | 21.637 | -3.563 | 3.66E-04** |
|  | Males | 953 | -59.841 | 30.515 | -1.961 | 0.050* |
|  | Females | 1356 | -57.632 | 35.135 | -1.640 | 0.101 |
| Age at first intercourse | Sexes pooled | 6107 | 82.351 | 13.501 | 6.099 | 1.07E-09** |
|  | Males | 2609 | 61.357 | 18.277 | 3.357 | 0.001** |
|  | Females | 3498 | 118.068 | 21.330 | 5.535 | 3.11E-08** |
| Asexuality (0=no, 1=yes) | Sexes pooled | 8719 | -0.249 | 0.624 | -0.398 | 0.690 |
|  | Males | 3688 | 0.041 | 0.893 | 0.046 | 0.963 |
|  | Females | 5031 | -0.412 | 1.046 | -0.394 | 0.693 |
| Sociosexuality | Sexes pooled | 1982 | 8.057 | 20.799 | 0.387 | 0.698 |
|  | Males | 738 | 1.667 | 35.656 | 0.047 | 0.963 |
|  | Females | 1244 | 31.000 | 31.675 | 0.979 | 0.328 |
| Self-reported physical attractiveness | Sexes pooled | 1938 | -8.810 | 23.563 | -0.374 | 0.708 |
|  | Males | 714 | -27.080 | 38.964 | -0.695 | 0.487 |
|  | Females | 1244 | 30.095 | 34.882 | 0.863 | 0.388 |
| Observer-rated facial attractiveness | Sexes pooled | 1354 | 26.993 | 31.723 | 0.851 | 0.395 |
|  | Males | 624 | -59.874 | 40.013 | -1.496 | 0.135 |
|  | Females | 730 | 31.430 | 47.254 | 0.665 | 0.506 |
| Romantically desirable | Sexes pooled | 1945 | 4.305 | 21.894 | 0.197 | 0.844 |
|  | Males | 719 | 14.664 | 35.000 | 0.419 | 0.675 |
|  | Females | 1226 | -0.094 | 32.210 | -0.003 | 0.998 |

\*Nominally significant association ( $p < 0.05$ ). \*\* Significant association after Bonferroni correction for multiple testing (0.05/11). SE; standard error of the slope estimate.

### Genetic overlap with complex traits and disease risk

We computed the genetic correlations of sexlessness with a wide range of traits using LD Score regression^15^. Genetic correlations measure the extent to which genetic effects are shared with other traits. The genetic correlations between sexlessness and childlessness (among those who had had sex) were 0.68 (SE = 0.10, *p* = 7.0E-11) for females and 0.65 (SE = 0.09, *p* = 7.0E-14) for males, indicating substantial genetic overlap. Most notably among other traits, there is a pattern of strong positive genetic associations of sexlessness with indices of cognitive ability and SES (Figure 6a, Table S4). We also found substantial negative genetic correlations between sexlessness and substance use phenotypes, with genes underlying sexlessness overlapping with those associated with less substance use. In addition, we found positive genetic correlations with autism and anorexia and negative correlations with ADHD and posttraumatic stress disorder and a range of phenotypes related to personality and social connection (extraversion, relationships with friends and family etc). A visualisation of the genetic correlation pattern between all traits is presented in Figure 6b. The graph shows that sexlessness for males and females are in the same cluster and that they both cluster strongly with a wide variety of traits, in particular traits related to SES and substance use. The genetic correlations of childlessness with the same selection of traits (Figure S3) show a highly similar pattern of correlations.

**Figure 6.**
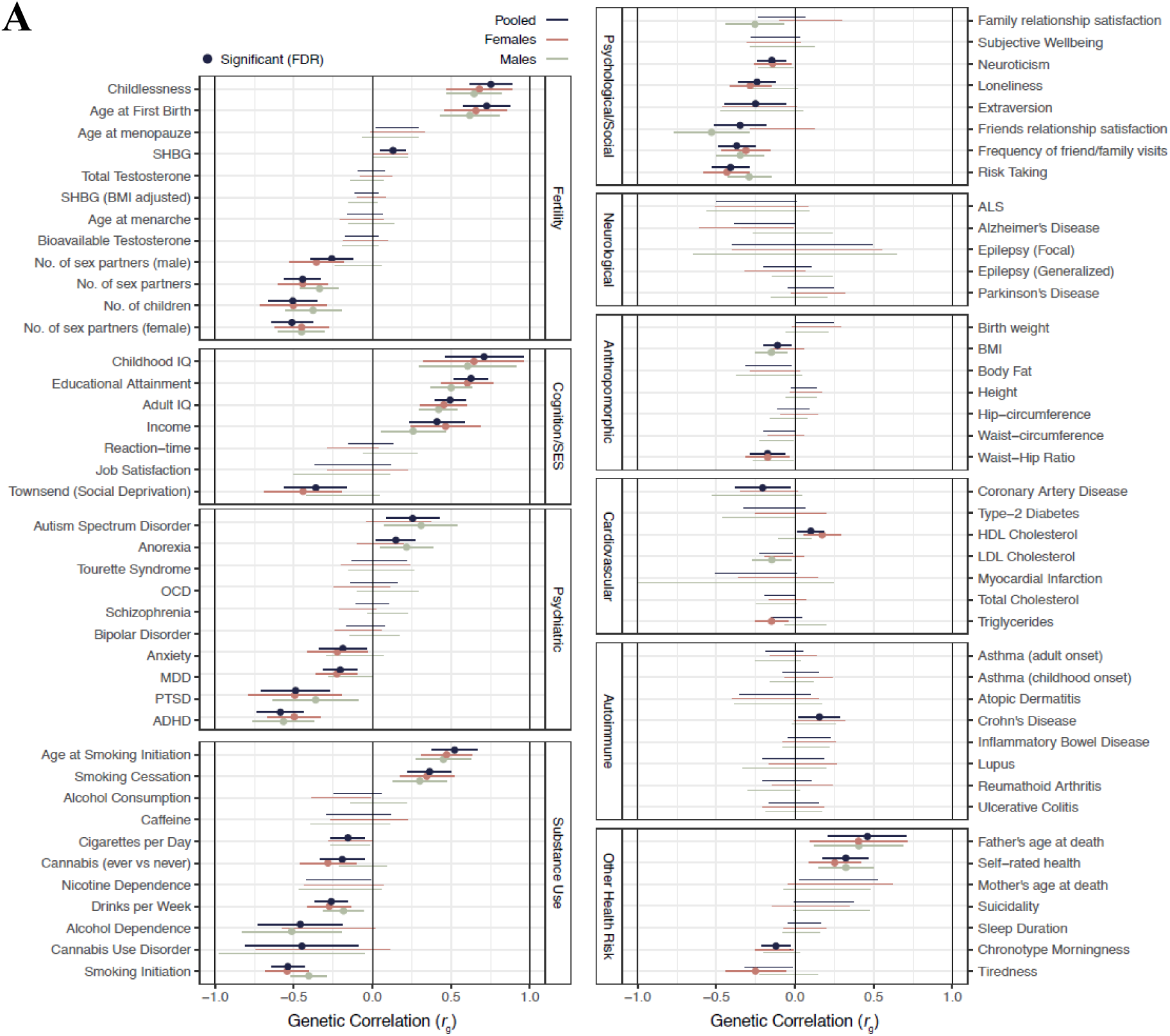
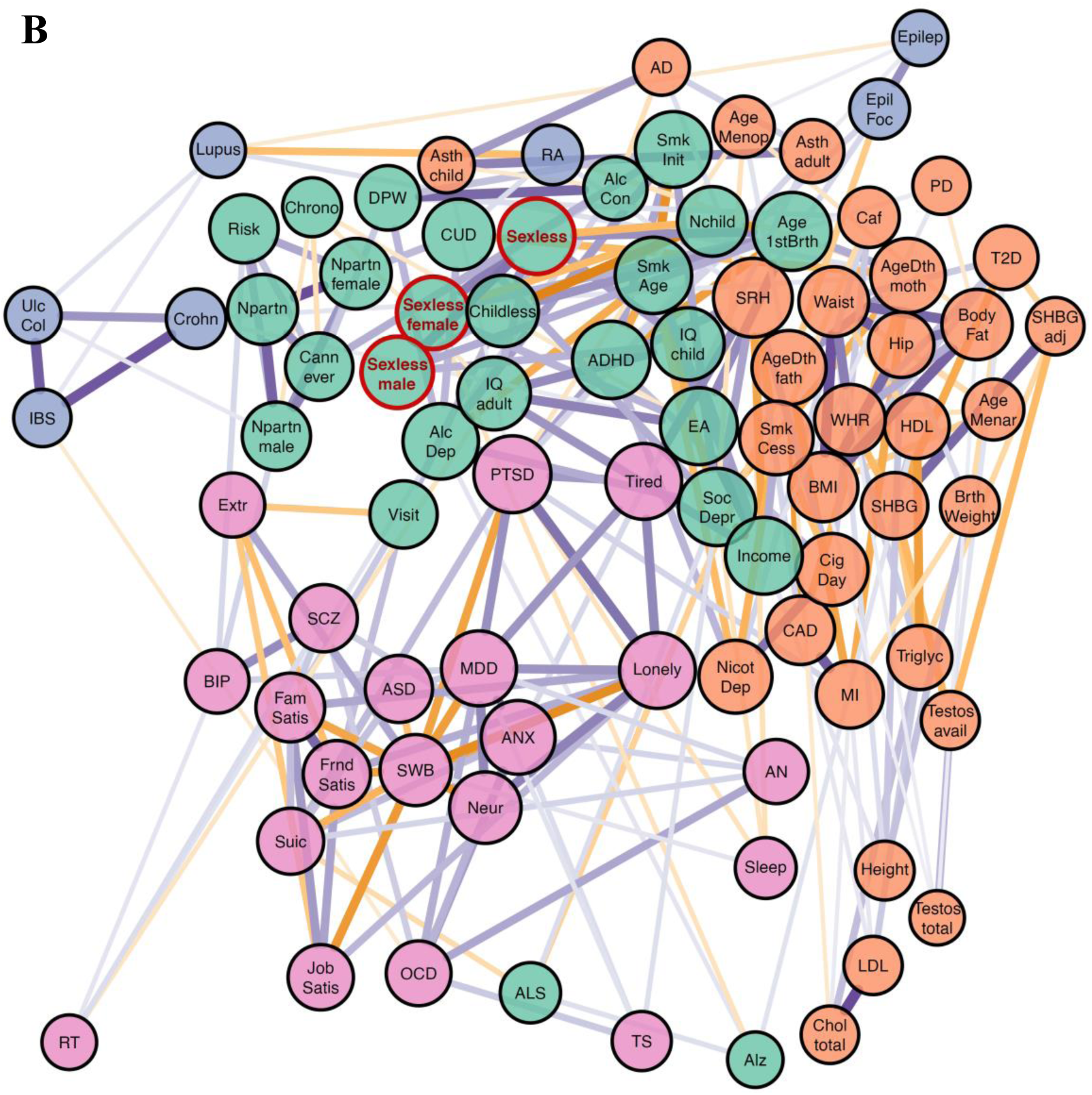
(below): Genetic overlap between sexlessness and a variety of complex traits. Panel **A**; Genetic correlations (rg) between sexlessness and a wide variety of complex traits computed with LDSC regression. Panel **B**; Visualization of the SNP-based genetic correlations between sexlessness and all other traits as a graph. Each vertex (node) represents a trait, with the colouring denoting different trait groups). Weighted undirected graphs were created with the absolute genetic correlation as connection strength (represented by line thickness). Orange edge colours represent negative genetic correlations and purple edges positive ones. Vertex size is based on eigenvector centrality with a minimum offset. Vertex layout was based on the Fr<u>ü</u>chterman and Reingold algorithm^16^. Colouring is based on a clustering algorithm on the genetic correlation matrix, and reflects what cluster the sexlessness belongs to. Phenotype names corresponding to the abbreviations can be found in Table S4.

## Discussion

We explored the correlates of sexlessness (never having had sex) by estimating phenotypic and genetic correlations with other traits and associations of variation in sexlessness rates and sex ratios and income inequality across regions of birth and residence. Prior knowledge about the correlates of sexlessness was limited. Our results confirm findings from earlier studies that sexless individuals tend to be more educated and less likely to use alcohol and drugs^7^. We also found direct evidence that sexless men are more likely to live in regions with a higher proportion of men as compared to women and in regions with greater income inequality, strengthening previous indirect evidence of such effects based on geographic localisation of Twitter posts that use incel language^9^. Beyond strengthening these previous findings, we also reported a wealth of other associations, at both the phenotypic and genetic levels, as well as sex differences in these associations and comparisons to corresponding correlations for childlessness.

Phenotypically, there were notable associations with indicators of poor mental wellbeing, especially for men. Sexless men tended to suffer more from nerves, unhappiness, loneliness, and were less likely to believe their life is meaningful. Throughout this discussion, the issue of causality is ever-present – the causal pathways underlying these cross-sectional associations are likely to be complex. In the case of the affective wellbeing indicators, one possibility is that never having had sex takes a toll on one’s happiness, given that sex is a fundamental human drive, deeply important in our evolutionary history. Other results in our data point to indirect routes in the same direction – sexless individuals (especially men) were much more likely to live alone, have less frequent social visits, and lack a confiding relationship or anyone to confide in. There were corresponding genetic correlations with these social variables too. It seems likely that sexual partners (who are often intimate emotional partners too) can provide strong social support, and lacking any such partner could be detrimental for mental wellbeing. But it may also be that poor mental wellbeing makes approaching or attracting potential sexual partners more difficult, and third variables could increase the likelihood of both sexlessness and poor mental wellbeing (and social support). One possibility in this latter vein is that the same genetic variants predispose to both sexlessness and poor mental wellbeing. But the genetic correlation of sexlessness suggest this possibility is unlikely with regard to the affective traits in question – the genetic correlations of sexlessness with major depressive disorder and anxiety were negative, not positive. On the other hand, autistic spectrum disorder showed a substantial positive genetic correlation with sexlessness, consistent with the interpersonal difficulties characteristic of the spectrum; autistic individuals commonly report difficulties developing intimate relationships^17^.

There were also phenotypic associations of sexlessness with measures related to lower physical robustness, including low grip strength and lean arm mass, and an earlier age of wearing glasses. The strength-related variables were only related to sexlessness in men, which is consistent with previous evidence that physically weaker men are less able to attract sexual partners^18^, whereas strength does not relate strongly to women’s sexual appeal. Another possibility might be that low testosterone leads to physical weakness as well as low sex drive – but testosterone measures did not show substantive associations with sexlessness at either the phenotypic or genetic level. Regarding the association with having worn glasses from an early age, focus groups have reported negative experiences from wearing glasses as children, being called “nerds” and “geeks” and generally being regarded as unattractive^19^. Glasses worn by adults seem not to be regarded as unattractive in general^20^; it could be that wearing glasses at an early age disrupts early adolescent dating experiences, which in turn impacts later sexual success^21^. Again, there may be other causal explanations, but they are not as obvious in this case.

Phenotypic and genetic correlations revealed links of sexlessness with less use of alcohol, cigarettes, and cannabis. It is unclear to what extent these links are underlain by risk aversion (risk-taking was negatively associated with sexlessness, genetically and phenotypically), less orientation towards seeking pleasure, or higher general inhibition. It could also be that taking alcohol and drugs directly reduces inhibitions that might otherwise preclude sexual activity. Alternatively, third variables could be involved. We found a genetic correlation of sexlessness with low extraversion – perhaps introverts are less likely to be in contexts (e.g. parties) where alcohol and drugs are involved, and are also less likely engage in social situations that facilitate meeting potential sexual partners.

Previous work has linked lifetime sexlessness with higher education significantly only in women^7^, though other work has shown periods of sexlessness, or sexlessness in young people, to be associated with lower education and lower income^6^. Here we show a small but highly significant positive phenotypic association of sexlessness with education in both men and women; we also found a negative association with household income and a positive association with the economic deprivation (Townsend index) of their neighbourhood, but these are likely reflect the economic ramifications of remaining single (e.g. household income will be lower than for couples). Also note that the genetic correlations of sexlessness with these traits are actually in the opposite direction of the phenotypic correlations.

More striking are the substantial (∼0.5) genetic correlations not only with higher education but with higher childhood and adult intelligence (IQ), as well as higher income and socioeconomic status. On the face of it, these associations are counterintuitive, especially from an evolutionary perspective in which intelligence and resources are supposed to be attractive traits in a potential partner, though see Driebe et al. (2021)^22^. In line with that perspective, a recent study also using UK Biobank data showed that men who carry a higher burden of rare deleterious genetic variants (that ablate protein-coding genes) have a lower educational attainment and IQ and fewer children. Moreover, the association between the burden of deleterious variants and number of children was partly mediated by sexlessness^23^. Obvious explanations for the positive associations we found, which apply to both men and women, are not apparent to us. We might wonder whether conscientiousness facilitates commitment to education or religion at the expense of seeking sexual partners. Still, it is difficult to explain the even higher estimates of genetic correlations with childhood IQ, which is supposedly less strongly related to conscientiousness than is educational attainment.

In making sense of the observed pattern of correlates of sexlessness, it would be remiss to overlook the resemblance of this pattern of characteristics – introverted, wearing glasses at a young age, intelligent, academically successful, physically weaker, socially disconnected, lonely, higher on the autistic spectrum, nervous, engaging less with drugs and alcohol – to the stereotype of a “nerd”, which is in turn associated with lack of romantic success^24^ (indeed, being unattractive is in several dictionary definitions for “nerd” ^25^). The nerd stereotype is perhaps most associated with adolescence, though, whereas the participants in this study are between 39 and 73 years old. It is worth noting again that adolescent experiences (or lack thereof) and identity formation may in some cases have long-lasting effects with regard to sexlessness^21^.

The preceding characterisation implies that sexlessness is primarily driven by difficulties obtaining partners, but could the observed correlates of sexlessness be better explained by lack of sexual attraction (i.e. asexuality)? Asexuals comprise around 1% of the population, similar to our late-life virgins as a proportion of our sample, though more than half of asexual individuals in a large probability sample were not virgins3; personal communication. However, that large probability sample also revealed that, relative to the broader population, asexuals were on average less educated and lower in personal socioeconomic status^3^– so it does not seem that our pattern of genetic correlations could be straightforwardly explained by characteristics known to be associated with asexuality. We also found no association of asexuality in the Australian sample with our sexlessness polygenic score, but given the small number of asexual individuals there would have been very low power to detect any true association. Last, the association of male sexlessness with geographic regions with relatively fewer women and greater income inequality is hard to explain in terms of asexuality, but straightforward to explain in terms of difficulty obtaining partners see ^9^. Nonetheless, surely asexuality does contribute to the phenotype we capture; large-scale data on sexual attraction as well as sexual behaviour would be required to tease these components apart.

From the point of view of the potential utility of sexlessness in evolutionary analysis, there are several findings to note. First, sexlessness showed substantial negative genetic correlations with number of opposite-sex partners (for both males and females) and number of children, variables that have elsewhere been used as indicators of fitness. The genetic effects underlying sexlessness thus overlap to substantial degree with those underlying broader variation in reproductive and mating success, even though these latter two variables are themselves not positively genetically correlated^26^. As described in the introduction, sexlessness might be in some ways a ‘purer’ (though less statistically powerful) proxy for ancestral fitness than these other variables, in that it is less likely to be driven by variables less related to ancestral fitness, such as sociosexual orientation (especially in women), mate choosiness, family planning, career-orientation, and the like. (Note, though, that it is still subject to cultural changes; see Box 1). Comparing phenotypic and genetic correlates of sexlessness to those of childlessness, there are many similarities, with patterns of positive genetic correlation of both traits with indices of cognition and SES, autism, and anorexia and negative genetic correlations with substance use, ADHD, and a range of phenotypes related to personality and social connection. However, phenotypically, we show that sexless individuals differ from non-sexless individuals on more traits than childless versus non-childless individuals (note; we chose a cut- off of ΔR^2^ = 1%). A clear difference is that childlessness does not have the same associations with unhappiness and loneliness, which may relate to childlessness more commonly reflecting a free choice than is the case for sexlessness. There is substantial evidence that the drive to have sex is much more ubiquitous than the drive to have children, e.g. more than 20% do not want to have children in a representative sample from Michigan^27^. Also, sexlessness, but not childlessness, is associated with less substance use.

An example of how sexlessness could be used to strengthen evolutionary inferences pertains to the sexual selection hypothesis of human intelligence^28^, which proposes that our extraordinary intelligence evolved because of its contribution to individuals’ mating success (via female preference for smarter mates). Previous work had shown that intelligence was negatively correlated with number of children^29^ and uncorrelated with number of sexual partners^30^, but a proponent of the hypothesis could argue that these observations are not necessarily disconfirmatory. Regarding number of sexual partners, intelligence might attract higher quality mates rather than a greater quantity, and regarding number of children, more intelligent people might focus on professional careers at the expense of having children (which would not have been a factor in the evolutionary past). But these rebuttals could not explain away the genetic correlation of intelligence (and education and income) with sexlessness. Therefore, our findings strengthen the case against the sexual selection hypothesis of intelligence (along with other data indicating that high intelligence is not generally found attractive^22^).

A strength of our study was that we were able to capitalise on publicly available GWAS summary statistics, allowing estimates of genetic correlations of sexlessness with other traits that were not assessed in the UK Biobank itself. The genomic data also enabled examination of questions that are impossible to answer with phenotypic data only. For example, we found that male and female sexlessness were associated with overlapping but not identical genetic markers; similarly, the genetic markers associated with sexlessness overlapped with those associated with childlessness among people who had had sex. Nonetheless, genetic correlations computed from GWAS summary statistics must be interpreted with caution. Genetic correlations between complex traits can reflect different types of pleiotropy, but also arise through gene-environment correlations^13,31^. The fact that the sexlessness polygenic score correlated in the predicted direction with the related variables in an independent Australian sample gives us confidence that our GWAS results are tapping into real variance in sexlessness and not only structural artifacts of the UK population. In addition, our UK-Biobank within- versus between-family polygenic score analysis revealed that the GWAS signal only captures gene-environment correlation effects to a very small degree, at least in terms of environments that differ between families. Still, as already mentioned, there is much uncertainty about the causal processes underlying the observed genetic (and phenotypic) correlations, and this limitation must be kept in mind when interpreting the findings. Research using different methodologies and different populations may help to triangulate a deeper causal understanding of the social and biological underpinnings of sexlessness.

## Methods

The primary analyses (GWAS and analyses of phenotypic and environmental correlates) were carried out in the UK-Biobank sample. Subsequently, polygenic score analyses were performed in an independent target sample from Australia; this sample is described below in the section on ‘polygenic score analyses’.

### Participants

Participants were from the UK Biobank^10^, a large-scale prospective study with longitudinal follow-up, which contains genetic and health information from half a million participants from the UK. Participants were recruited between 2006 and 2010 across 22 assessment centers throughout the UK and were between 39 and 73 years old at wave 1. Data on health, cognition and lifestyle as well as blood, urine and saliva samples were collected. We analyzed data from 455,978 individuals (247,340 females, 208,638 males, 1 gender missing) with European ancestry for whom genetic and phenotypic data were available. UK Biobank has received ethical approval from the National Health Service North West Centre for Research Ethics Committee (reference:11/NW/0382).

### Phenotypes

Data on sexlessness (data-field 2139) were available in the UK Biobank sample for 405,117 individuals (218,744 females and 186,373 males). Of those, 3,929 individuals (2,068 females and 1,861 males, both ∼1%) responded “never had sex” to the question “What was your age when you first had sexual intercourse? (Sexual intercourse includes vaginal, oral or anal intercourse)”. For our analyses sexlessness was coded as 1 for individuals who never had had sex and 0 for participants who have had sex.

Data on childlessness (data-fields 2734 and 2405 for females and males) were available for 455,979 individuals, of which 44,118 males reported to have not fathered any children and 46,251 females reported haven given birth to zero children (Ntotal = 90,369). For our analyses childlessness was coded as 1 for individuals who never had a child and 0 for participants who have had a child. We excluded individuals that have not had sex (3,929) from the analyses focused on childlessness (remaining sample 452,050 individuals). Note, a small subset of participants (N=24) reported they had children, but did not have sexual intercourse in their life.

To examine phenotypic correlates of sexlessness, we selected 253 health, psychological, and behavioral phenotypes from the survey data from UK Biobank; these phenotypes were broadly related to domains of mental health, sleep, exercise, substance use, risk taking behavior, cognition, health, and occupation. Selected phenotypes had a sample size above N=10,000 (for binary traits we used effective sample size: Neff = 4/(1/Ncases + 1/Ncontrols). For an overview of all included traits, see Table S1. We used the first available measurement for each individual; if data were not available at the first wave, we took the second, otherwise the third, etc. For continuous phenotypes, theoretically implausible values were set at missing and otherwise data were winsorized at 4 standard deviations (SDs) from the mean. The continuous (including ordinal) phenotypes were then standardized with a mean of 0 and a SD of 1. No changes were made to binary phenotypes.

### Statistical analyses

#### Phenotypic correlates of sexlessness and childlessness

We performed 249 logistic regression analyses^i^, regressing sexlessness on each of the selected health, psychological, and behavioural phenotypes, while including age, sex, age-squared, age-by-sex interaction and year of birth as covariates. We performed another 251 logistic regression analyses, regressing childlessness on each of the selected health, psychological, and behavioural phenotypes, including the same set of covariates. To estimate the effect size for each phenotype separately, we subtracted the total explained variance of the model without the respective phenotype (with covariates) from the total variance explained of the full model (including the phenotype and covariates). Significant phenotypes (Bonferroni corrected *P*- value<0.05) that explained ≥1% of the variance or more were considered to be relevant correlates of sexlessness or childlessness. Analyses were repeated for females and males separately. Analyses were performed in R-studio version 4.0.3.

#### Sexlessness in relation to regional sex ratio and regional income inequality

We investigated whether there was an association of sexlessness and the sex ratio of the region in which people were born and the region they currently lived, and between sexlessness and income inequality in these regions. The regional sex ratios (% women) at the Middle Layer Super Output Area (MSOA) level were estimated from the census carried out by the Office of National Statistics in 2011 and obtained from the Nomis website (https://www.nomisweb.co.uk/sources/census_2011). Birth place locations of the UK Biobank participants were based on the rounded coordinates from UK Biobank fields 129 (latitude) and 130 (longitude), and current address locations were based on the rounded coordinates in the UK Biobank fields 22702 (longitude) and 22703 (latitude). The process of linking UK Biobank participants to their MSOA region is described in more detail in Abdellaoui et al. (2022)^31^.

Income inequality was determined using the Gini coefficient, a commonly used measure of income inequality within a population^32^. It quantifies the dispersion of income among individuals or households, providing a summary statistic of the income distribution. A Gini coefficient of 0 represents perfect equality, where everyone has the same income, while a value of 1 represents complete inequality, where one individual or household possesses all the income.

The Gini coefficient was computed using the UK Biobank data, containing information about household income of individuals and their respective MSOA regions. Household income was categorized as follows: 1) Less than £18,000, 2) £18,000 to £30,999, 3) £31,000 to £51,999, 4) £52,000 to £100,000, 5) Greater than £100,000. Mid-point values were assigned to each income category, except for the open-ended category “Greater than £100,000,” where an arbitrary value of £125,000 was assigned. The Gini coefficient was computed for each MSOA based on the recoded income values. The data were grouped by MSOA, and the Gini coefficient was computed for each region separately. MSOAs with only one observation were excluded from the analysis to ensure statistical robustness. The Gini coefficient was calculated using the *ineq* function from the R package *ineq*, based on the grouped income data.

#### Genome-Wide Association Study (GWAS) of sexlessness

A GWAS was run in fastGWA^33^ on 10.6 million single nucleotide polymorphisms (SNPs) in 404,470 participants using a linear mixed model approach correcting for the genetic relatedness matrix (GRM) and 25 genetic PCs to control for cryptic relatedness and population stratification, respectively. The GWAS was performed for participants with European genetic ancestry only. Ancestry was determined by two rounds of principal component analyses. For the first round, the UKB dataset was projected onto the first two principal components (PCs) from the 2,504 participants of the 1000 Genomes Project. Based on these PCs, a total of 456,064 participants from UKB were identified to have a European ancestry, of which 400,573 had phenotypic data on sexlessness. A second PCA was run on these 456,064 participants to capture British/European ancestry differences. The first 25 PCs of this second PCA were used as covariates in the GWAS to control for population stratification. More details on genotyping, QC, and PCAs are described in Abdellaoui et al. (2022)^31^. We also performed GWASs separately for males (N=186,035) and females (N=218,435). We used the GWAS summary statistics to compute gene-based *P*-values in MAGMA^34^ for 18,714 protein-coding genes using FUMA^35^.

#### Polygenic score analyses

We constructed polygenic scores for sexlessness from the GWAS findings to validate their ability to predict sexlessness and related traits in independent samples, and to examine gene-environment correlations in the GWAS signal. Polygenic scores are measures of an individual’s genetic predisposition for a trait, based on the sum of alleles weighted by their estimated effect on the trait of interest (here sexlessness).

GWAS signals can capture gene-environments correlations. In particular, previous studies have shown SES-related traits have strong levels of gene-environment correlations, which get captured by GWAS signals and polygenic scores. Because of these gene-environment correlations, polygenic scores for SES-related traits do a better job predicting between-family differences than within-family differences. To test the presence of gene-environment correlations on the family-level for sexlessness, we predicted sexlessness within and between families in the UK-Biobank sample with the same approach as Selzam et al. (2019)^14^.

Polygenic score analyses were performed in the UK-Biobank sample and in an Australian sample.

##### UK-Biobank

As polygenic scores should be calculated in an independent sample, we first repeated the GWAS on sexlessness in a subset of the sample, excluding all siblings (and their relatives) and then built polygenic scores for the siblings. The PGSs for sexlessness, the genome-wide sum of alleles weighted by their estimated effect sizes, were computed using the SBLUP approach^36^, which maximizes predictive power by creating scores with best linear unbiased predictor (BLUP) properties, while accounting for linkage disequilibrium (LD). We used a random sample of 10,000 unrelated individuals from UK Biobank that were imputed using the Haplotype Reference Consortium (HRC) reference panel as the LD reference sample. For more details on the GWAS excluding siblings and polygenic score computation, see Abdellaoui et al. (2022)^31^. We then fitted the following models in 19,310 sibling pairs (of which 478 individuals never had sex):

**model 1:** The model, with the polygenic score on an individual-level as fixed effect:

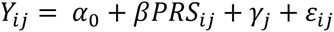

**model 2:** The model with between-family fixed effects added:

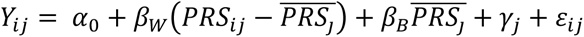

The *β* from model 1 was .016 (SE = .005, *p* = .002). In model 2, the *βW* was .015 (SE = .01, *p* = .14) and the *βB* was .016 (SE = .006, *p* = .007). Individual-level effects decreased by 7.4% when adding between-family effects.

##### Australian twin sample

To further validate the GWAS findings, we also examined the associations between polygenic scores for sexlessness and a number of relevant traits related to sexuality, mating, and attractiveness in an independent Australian sample. The target sample comprised individuals from the Australian Twin Registry (ATR) and the Queensland Twin Registry (QTwin) (both from QIMR Berghofer, Australia) for whom both genotypic and phenotypic data were available (N ranged between 1,354 and 13,532)^37–40^. Individuals participated in various studies between 1988 and 2018. Outcome variables included 11 items about romantic partners, sexual partners, age at first intercourse, asexuality, sociosexuality (orientation toward uncommitted sexual relationships), physical attractiveness, and romantic desirability (see Table S6 for information about the variables included).

The PGSs for sexlessness were computed using the SBLUP (see above). Generalized estimating equation (GEE) modelling was applied to test whether the PGSs for sexlessness predicted the outcome phenotypes in the target cohort with family number as cluster variable. An ‘independence’ covariance matrix was used to model family relatedness and tests were based on robust (sandwich-corrected) standard errors.

Age, age-squared, birth year, ten genetic PCs, and sex, age-by-sex interaction (sex-pooled version only) were included as covariates in the model plus a binary variable to account for genotyping platform effects (i.e. imputed from HapMap-derived genotyping chips, vs imputed from 1000 Genomes-derived chips); and (when applicable) a binary variable to slight difference in response coding between two groups of similarly-worded questionnaires. Analyses were performed in R version 3.6.2. PGS analyses were also performed separately for males and females.

#### Genetic correlation analyses to examine genetic overlap with complex traits and disease risk

We estimated genetic correlations of sexlessness with a range of health, psychological, and behavioural phenotypes (N=82 traits, Table S4 (and see Table S5 for results for childlessness)). Genetic correlations were computed with LD score regression^11,14^, which estimates the slope from the regression of the product of z-scores from two GWASs on the LD score, reflecting the genetic covariation between two traits based on all polygenic effects captured by the included SNPs. These analyses were done on the ∼1.3 million genome-wide HapMap SNPs used in the original LD score regression studies^11,14^. The LD information used by these methods was based on data from European populations from the HapMap 3 reference panel. Genome-wide genetic correlations were estimated for (1) sexlessness with childlessness, (2) sexlessness (males) with sexlessness (females), and (3) sexlessness (and childlessness) with a range of health, psychological, and behavioural phenotypes. Table S7 contains a list of the GWASs that were used to compute genetic correlations with sexlessness.

To visualise the clustering based on genetic correlations between sexlessness and other traits, we used graph analysis on the absolute genetic correlation matrices. R package iGraph (v1.3.1)^41^ to visualize the connectivity strength based with the vertex (node) size representing eigenvector centrality of the trait^42^ and edge (correlation) strength represented by hue and width. To avoid clutter, edge plotting was restricted to the four strongest edges. Vertex layout was based on the 2D Fruchterman-Reingold algorithm^15^. Vertex colour identifies grouping based on Louvain clustering^43^.

## Data Availability

Code is available at https://osf.io/6h7xj/?view_only=57a7810bd4034fe78812ce70ee4880d0, and the GWAS summary statistics will be made available at GWAS Catalog upon publication (www.ebi.ac.uk/gwas/downloads/summary-statistics).
The UK Biobank data is available upon request via: https://www.ukbiobank.ac.uk/enable-your-research/apply-for-access

## Acknowledgements

This study was conducted using UK Biobank resources under application number 40310. K.J.H.V. and A.A. are supported by the Foundation Volksbond Rotterdam. A.A. is supported by Amsterdam UMC / UvA Talent Fellowship D&I. R.A is supported by the Czech Health Research Council (grant nr. NU21J-04-00024) and a MSCA Fellowships CZ, OP JAK (grant nr. CZ.02.01.01/00/22_010/0002828).

## Author Contributions

Research idea/design: AA, LWW, BPZ, KJHV; Analysed data: AA, LWW, SDG, DJAS; Wrote the paper: AA, LWW, BPZ, KJHV; Critical review of paper for important intellectual content: JAP, DJAS, RA, NGM, FU, MAM.

## Data and materials availability

Code is available at https://osf.io/6h7xj/?view_only=57a7810bd4034fe78812ce70ee4880d0, and the GWAS summary statistics will be made available at GWAS Catalog upon publication (www.ebi.ac.uk/gwas/downloads/summary-statistics).

## Supplementary Figures

**Figure S1:**
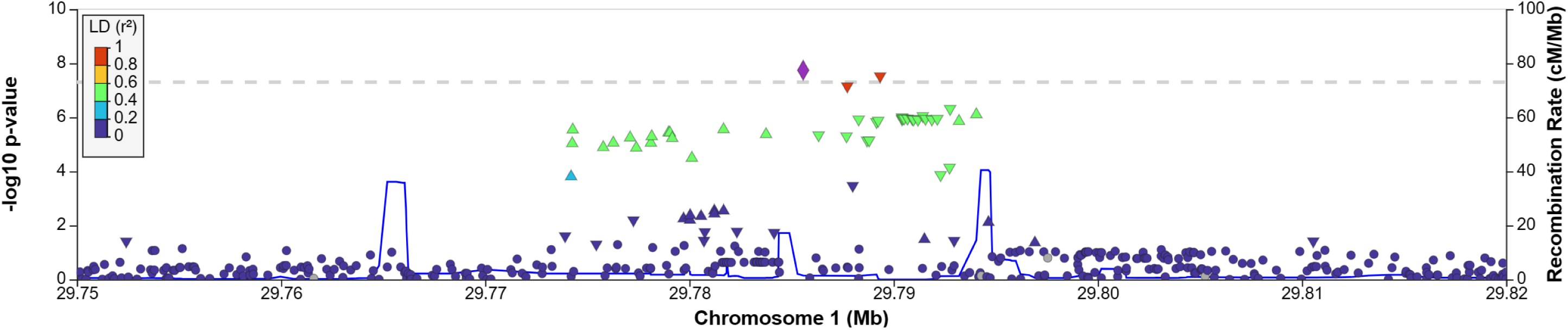
Locus zoom plot of the region with the genome-wide significant SNP.

**Figure S2:**
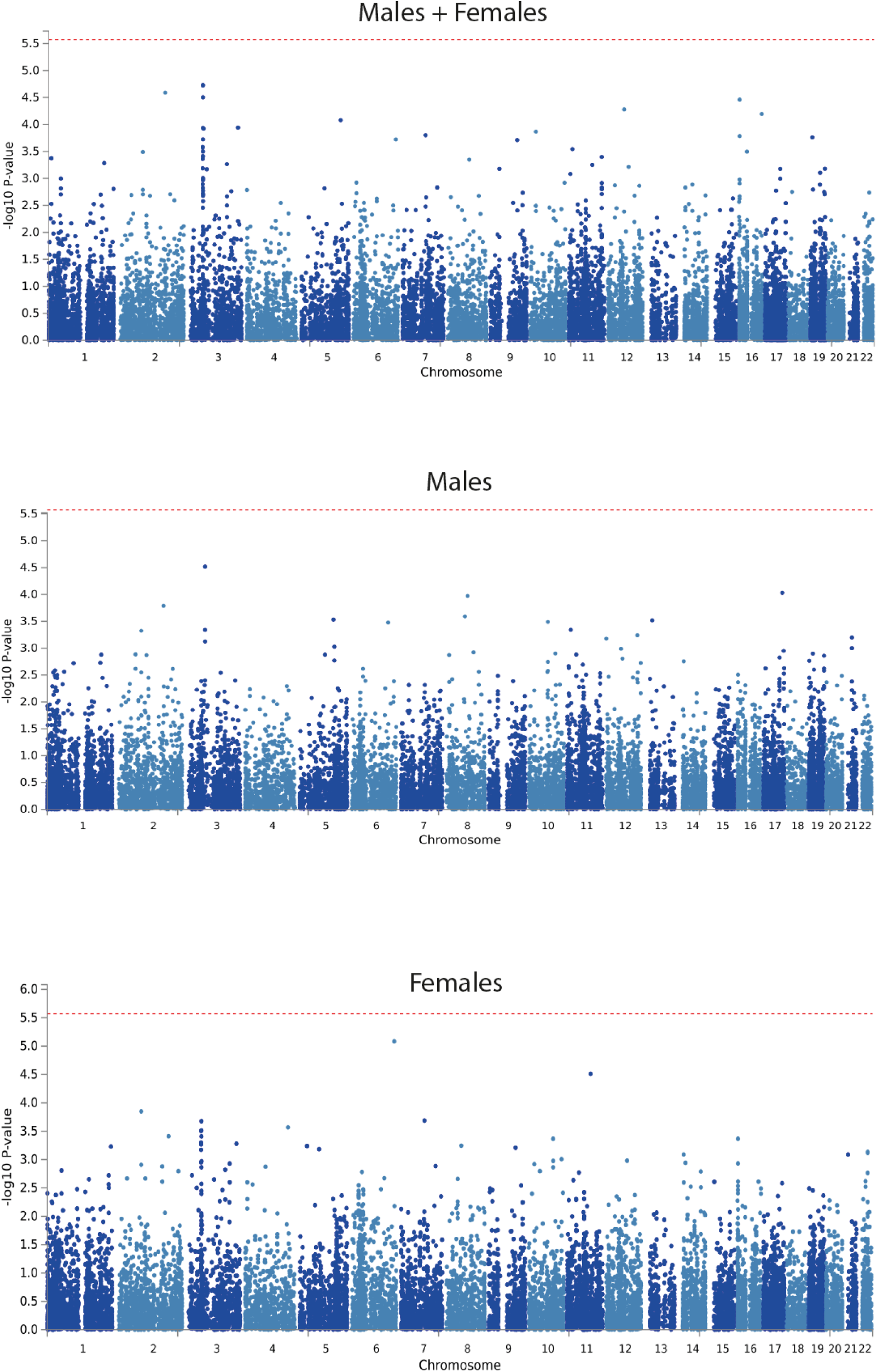
Manhattan plots of the MAGMA gene-based tests for males and females pooled and separately. The red line indicates the threshold of genome-wide significance.

**Figure S3:**
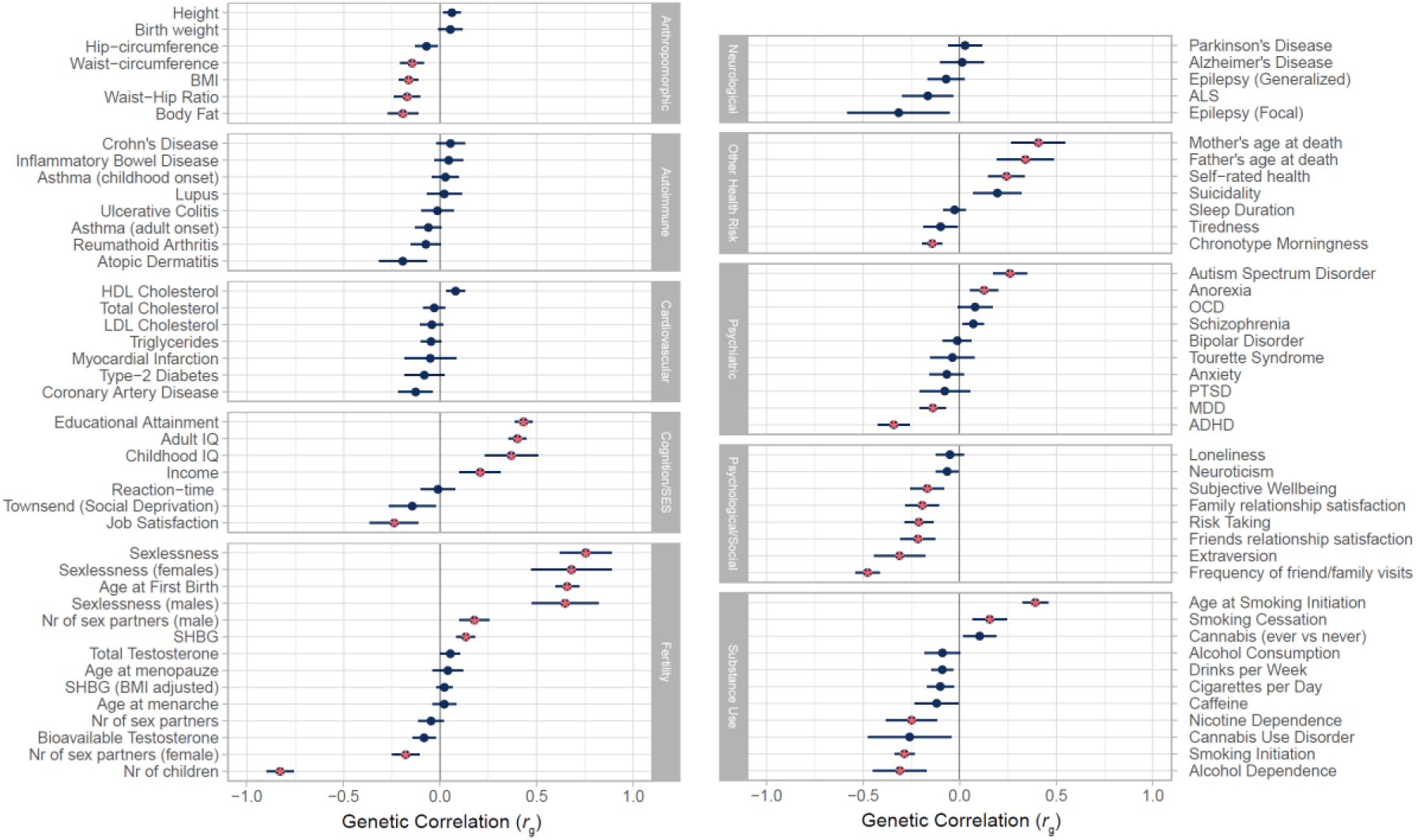
Genetic correlations (rg) between childlessness and a wide variety of complex traits, computed with LDSC regression.

## Footnotes

i Note that the number of analysed phenotypes differs between the analyses for sexlessness (249) and childlessness (251), as some survey items were not asked to people who reported to have never had sexual intercourse and some not to people who have no children.

